# Integrating stimulus-organism-response model and theory of planned behavior to explore athletes’ intention to receive the COVID-19 vaccine booster—A moderated mediation model

**DOI:** 10.1101/2023.11.13.23298480

**Authors:** Wenpeng Zhan, Qianting Deng, Van Bac Nguyen, Tran Phan Duc Anh, Phan Danh Na, An-Shin Shia, Gordon Chih-Ming Ku

## Abstract

This study aims to investigate the factors influencing athletes’ intention to receive the COVID-19 vaccine booster in Mainland China by integrating the stimulus-organization-response (SOR) model and theory of planned behavior (TPB) as the theoretical framework. Purposive sampling was used to select respondents from the National Games of the People’s Republic of China. Hard-copy questionnaires were utilized to collect data, resulting in 981 valid responses. Descriptive analysis and partial least squares structural equation modeling were used to analyze the data. The findings reveal that athletes’ subjective norm and knowledge significantly influence attitude, commitment, and perceived behavioral control. Attitude, commitment, and perceived behavioral control are verified as full mediators between subjective norm, knowledge, and intention to receive the COVID-19 vaccine booster. Knowledge to commitment is the most powerful path to predict athletes’ intention to receive the COVID-19 vaccine booster. Motivation moderates the relationships between knowledge, attitude, commitment, and perceived behavioral control. The integrating model’s explanatory power is 83.2%. Athletes’ knowledge is crucial in shaping a positive attitude, commitment, and perceived control, enhancing their intention to get the COVID-19 vaccine booster.

## 1. Introduction

The global sports community faced unprecedented challenges during the COVID-19 pandemic. Many sports competitions and training activities were halted due to anti-epidemic policies and regional lockdowns to prevent the virus’s spread. This situation significantly impacted athletes’ rights in Mainland China. Numerous important sporting events have been postponed or suspended, including high-profile ones like the Winter Olympics, the Chinese Basketball Association (CBA) Basketball League, and the Chinese Super League [1]. These disruptions severely affected the financial stability of sports organizations, the development of sports infrastructure, and, most importantly, athletes’ mental and emotional well-being [2, 3]. Although COVID-19 vaccination has eased these difficulties, it is essential to note that vaccine efficacy is limited, and the virus’s rapid mutations raise the risk of future outbreaks [4]. To ensure the health and safety of athletes and create a safe environment, receiving the COVID-19 vaccine booster is imperative. It is not only crucial for the sports industry but also for athletes’ well-being.

Upon the COVID-19 vaccine, most studies focus on the factors influencing the first-time vaccination. Only a few studies have made significant efforts to explore the factors influencing people’s intention to receive COVID-19 vaccine booster. Understanding these factors is crucial for enhancing revaccinated acceptance, increasing revaccination rates, and reducing the risk of infection and severe illness [4–9]. For example, Kwok, Li (4) found that older age, smoking, experiencing no adverse effects from a previous COVID-19 vaccination, greater confidence in vaccines and collective responsibility, and fewer barriers in accessing vaccination services are associated with higher intention of receiving vaccine booster. Liu, Lu (5) study verified that expectation confirmation toward receiving the COVID-19 vaccine affects the vaccination intention of COVID-19 vaccine boosters. The studies indicate that factors influencing the acceptance of COVID-19 vaccine boosters vary among different populations, such as healthcare workers, adults, etc., and individuals of different nationalities, such as Iran, Australia, etc. Consequently, each study presents various strategies to enhance the acceptance of COVID-19 vaccine boosters. However, no empirical research addresses the athletes’ acceptance of COVID-19 vaccine boosters.

The Theory of Planned Behavior (TPB) has been widely utilized to interpret the intention of vaccination across various contexts [10]. It has been applied to understand the intention to vaccinate against H1N1 [11], HPV vaccination [12], whooping cough vaccination [13][15], MMR vaccination [14], and hepatitis A vaccination [15]. However, while the TPB has proven to be a robust framework for explaining and predicting vaccination behavior [16], it has limitations in fully capturing behavioral intentions. As Catalano, Knowlden (17) have pointed out that TPB provides a valuable but partial explanation for the intention of vaccination. Therefore, there is a need to incorporate other models and variables into TPB to create a more comprehensive theoretical framework.

Stimulus-Organism-Response (SOR) is another well-known model widely used to identify the factors that trigger people’s behaviors. It assumes that an individual’s behavior is influenced by external stimuli affecting the mind, thereby eliciting specific actions [18]. Particularly, the SOR model has been applied to understand various behaviors in the COVID-19 context, such as purchasing behavior [19], information-avoidance behavior [20], seeking, passing, and providing COVID-19 information [21], travel intention [22], and COVID-19 vaccination [23]. While the SOR model offers a concise and robust theoretical foundation, combining it with the TPB theory can provide a more comprehensive understanding of individuals’ behavior [24, 25]. For example, some studies have found that subject norm does not significantly affect behavioral intention, necessitating additional theories or variables to explain this phenomenon [26]. Therefore, integrating the TPB and SOR model to explore factors influencing athletes’ intention to receive the COVID-19 vaccine booster, especially a lack of related studies, is essential to formulate effective strategies.

Reviewing the literature on vaccination in previous studies, knowledge [27–29] and commitment [30, 31] are identified as vital factors that affect individuals’ intention of vaccination. Knowledge is the most efficient power to stop the virus from spreading and protect oneself from being infected with COVID-19 [27]. On the other side, commitment ensures people can live in a healthy environment as COVID-19 continues to threaten our safety [31]. Thus, understanding the relationship between athletes’ COVID-19 vaccine knowledge, commitment, and intention to receive the COVID-19 vaccine booster helps formulate improved public health strategies. Moreover, motivation is a potential moderator to enhance the attitude toward vaccination [32]. Previous studies mentioned that people with a higher level of motivation usually have a more positive attitude toward vaccination than those with a lower level of motivation [32]. Therefore, motivation can potentially enhance athletes’ attitude toward the COVID-19 vaccine booster.

Understanding athletes’ intention to receive the COVID-19 vaccine booster is essential for a safe environment. Nonetheless, there needs to be more related studies. It is an urgent issue because there is hesitancy regarding COVID-19 vaccine boosters in China, especially among young individuals [33]. This situation may once again jeopardize athletes’ opportunities for competition and training. Developing strategies to enhance athletes’ intention to receive the COVID-19 vaccine booster is crucial in addressing the viral threat. Accordingly, the purpose of this study is to integrate the TPB and SOR model while incorporating two critical variables, knowledge and commitment, to examine their relationships to receiving the COVID-19 vaccine booster. This study also assesses the moderating effect of motivation in the model. The goal is to derive specific strategies from these findings for reference.

## 2. Literature Review

### 2.1 Theory of Planned Behavior and Stimulus-Organism-Response Model

The TPB is one of the popular frameworks for studying individual behaviors. It aids researchers in identifying the precursors to environmental behaviors and devising interventions to address these elements [34]. According to TPB, behavior is driven by personal attitude, subjective norm, and perceived behavioral control, which posits that behavior and norms influence three intention determinants and control beliefs, often called indirect predictors [35]. Within this theory, personal attitudes are characterized by individuals’ positive or negative evaluations of self-expression regarding a specific behavior [16]. Subjective norms and perceived behavioral control, on the other hand, pertain to an individual’s perception of social pressure and the ease of performing the behavior, respectively [16]. More favorable attitudes and subjective norms are associated with greater perceived behavioral control and an individual’s willingness to engage in the behavior [16].

Nevertheless, TPB still has limitations in explaining individual behavior, such as the person-factor issue [36]. Hill, Figueredo (36) argued that TPB focuses on individual cognitive factors and does not fully consider the broader environmental or contextual influences on behavior. Due to the dynamic nature of human behaviors, individuals might change their perspectives based on external stimuli, or personal experiences are ignored [37]. Therefore, athletes’ decisions regarding the COVID-19 vaccine booster might not be solely based on their attitudes or beliefs. External factors need to be considered [34].

The SOR model offers a theoretical perspective on how external stimuli influence individual behaviors through learned responses [38]. Rooted in existing research, this model proposes that when an individual encounters an external stimulus, it activates their internal emotional state. The Stimulus (S) stands for external triggers that initiate changes in one’s inner realm. The Organism (O) signifies the internal experiences tied to an individual’s cognitive and emotional processing. Notably, cognitive processing involves thought patterns based on the assimilation of information, while emotional processing pertains to feelings and emotions [39]. Lastly, the Response (R) encapsulates the resulting behavior that emanates from the interaction between the external stimulus and the internal state. This behavior directly manifests how the internal states influence individual reactions [40].

Integrating the TPB and SOR model can complement the limitations of explaining behavior within a single model. The SOR model provides a nuanced understanding of behavioral intention (intention to receive the COVID-19 vaccine booster) by emphasizing the emotional and affective responses (attitude, commitment, and perceived behavioral control) to external stimuli (knowledge and subjective norm), thereby addressing the gap in TPB’s cognitive-centric approach. Furthermore, the SOR framework offers a dynamic interplay between the environment and an individual’s internal processes, simplifying the operationalization and measurement of behavioral influences and embedding external environmental considerations. Accordingly, integrating the TPB and SOR offers a comprehensive framework combining cognitive and emotional determinants, enabling a more thorough understanding of athletes’ intention of the COVID-19 vaccine booster.

### 2.2 Research Framework and Hypotheses

#### 2.2.1 Stimulus: Subjective norm and Knowledge

Subjective norm can be regarded as an external stimulus. It refers to the influence of critical social individuals or groups and the pressure to conform to societal expectations, which can shape an individual’s behavioral intention [16, 41]. It reflects the individual’s perception of how significant others (e.g., family, friends, peers, or society) would like them to behave. Several studies have confirmed that subjective norm significantly affects individuals’ intention to receive the COVID-19 vaccine booster [42–44]. Since Athletes can internalize the beliefs of their social circles, it can further increase their willingness to get the COVID-19 vaccine booster.

Subjective norm affects behavioral intention and significantly affects individual emotional perceptions [45]. For example, an attitude towards a particular behavior is shaped by family, colleagues, friends, and experts, influencing buying behavior [46, 47]. Moreover, subjective norm shapes commitment by leveraging individuals’ innate desire for social acceptance and cohesion, with motivations ranging from seeking external validation and fearing social repercussions to desiring harmony and avoiding cognitive dissonance [48]. Especially when not receiving the COVID-19 vaccine booster could impact other athletes’ training and competitions, it might lead to heightened commitment. Accordingly, this study aims to investigate whether subjective norm can influence athletes’ attitude, commitment, and intention to receive the COVID-19 vaccine booster. The study proposes the following hypotheses:

H1: Subjective norm significantly influences athletes’ attitude toward COVID-19 vaccine booster.

H2: Subjective norm significantly influences athletes’ commitment of COVID-19 vaccine booster.

H3: Subjective norm significantly influences athletes’ intention to receive the COVID-19 vaccine booster.

Knowledge is the key to determinant the intention to receive the COVID-19 vaccine booster [49, 50]. Knowledge can equip individuals with accurate information, dispelling myths and fostering trust in the COVID-19 vaccine [49]. Sujarwoto and Maharani (50) mentioned that misinformation and less accurate knowledge are major contributors to COVID-19 vaccine hesitancy. Thus, accurate knowledge about the COVID-19 vaccines correlates with reduced hesitancy and increased acceptance. There are several studies proved that knowledge can positively enhance individuals’ attitude [51], commitment [52], and perceived behavioral control [53]. Consequently, it is plausible to hypothesize that the extent of athletes’ knowledge of the COVID-19 vaccine booster could significantly influence their attitudes and behaviors. The hypotheses are postulated as follows:

H4: Knowledge significantly influences athletes’ attitude toward COVID-19 vaccine booster.

H5: Knowledge significantly influences athletes’ commitment of COVID-19 vaccine booster.

H6: Knowledge significantly influences athletes’ perceived behavioral control of COVID-19 vaccine booster.

H7: Knowledge significantly influences athletes’ intention to receive the COVID-19 vaccine booster.

#### 2.2.2 Organism: Attitude, Commitment, and Perceived Behavioral Control

In the context of TPB, attitude is defined as the positive or negative psychological evaluation of engaging in a specific behavior [16], which substantially affects individuals’ behavioral intentions. It believes that people are more likely to engage in a behavior if they have a positive attitude, and vice versa [16]. Therefore, numerous studies have confirmed that attitude can enhance the intention to receive the COVID-19 vaccine booster [42, 53, 54]. For instance, Asmare, Abebe (53) found that attitude and subjective norm are statistically significant predictors of intention to receive the COVID-19 vaccine booster. Attitude is the strongest predictor in the model. Attitude shapes perception, and in the context of the COVID-19 vaccine booster, it is the pivotal force driving athletes’ intention to vaccinate, anchoring decisions in belief and sentiment.

Attitude in the expended TPB model can be regarded as a mediator. Attitude is a psychological response influenced by external stimuli, leading to positive or negative perspectives, and then resulting in specific behaviors [55–57]. Tarkar (56) verified that attitude is a vital mediator in connecting perceived benefits, perceived susceptibility, and intention to receive the COVID-19 vaccination. Accordingly, attitude within expended TPB is a vital link between individuals’ cognitive and emotional evaluations of their intentions to perform the behaviors. However, there is a lack of study on the COVID-19 vaccine booster issue from athletes’ perspectives. This study tests attitude’s role (direct and indirect effects) in the integrating model. The study proposes the following hypotheses:

H8: Athletes’ attitude significantly influence their intention to receive the COVID-19 vaccine booster.

H9a: Athletes’ attitude is a mediator between subjective norm and intention to receive the COVID-19 vaccine booster.

H9a: Athletes’ attitude is a mediator between knowledge and intention to receive the COVID-19 vaccine booster.

Commitment is widely acknowledged as an effective means of sustaining the value of entities or individuals in long-term relationships and generating desired behaviors [58]. Commitment is often associated with a strong emotional and psychological attachment to a particular behavior or outcome. It can enhance the strength of an individual’s intention toward a specific behavior [58, 59]. In vaccination studies, commitment is regarded as a causal factor that significantly drives actual vaccination behavior [59]. This assumption gains particular relevance in the context of COVID-19 vaccine booster, given the continuous mutation of the coronavirus and the limited duration of vaccine protection [60]. Therefore, this study considers commitment an extended variable and a mediator within the integrating framework, contributing to the comprehensive understanding of the relationship among athletes’ subjective norm, knowledge, and intention to receive the COVID-19 vaccine booster. The study proposes the following hypotheses:

H10: Athletes’ commitment significantly influences their intention to receive the COVID-19 vaccine booster.

H11a: Athletes’ commitment is a mediator between subjective norm and intention to receive the COVID-19 vaccine booster.

H11a: Athletes’ commitment is a mediator between knowledge and intention to receive the COVID-19 vaccine booster.

Perceived behavioral control is a crucial concept within the TPB as it pertains to an individual’s perception of their ability to control and carry out a specific behavior [16]. It reflects individuals’ assessment of the obstacles, challenges, and barriers they may encounter when executing the behavior. High perceived control can help overcome the barriers that make individuals believe they have significant control over the factors that facilitate the behavior [16]. This perception can significantly influence the likelihood of forming behavioral intentions and subsequently taking action [16]. Several studies have verified a causal relationship between perceived behavioral control and vaccination intention [61, 62]. For example, Liu, Wang (62) research demonstrated a significant correlation between women’s behavioral intentions regarding human papillomavirus vaccination and their perceived behavioral control. In the context of this study, perceived behavioral control refers to an athlete’s confidence in their ability to accept and undergo the COVID-19 vaccine booster. This study examines the effect of perceived behavioral control on athletes’ intentions to get vaccine booster against COVID-19. Therefore, the study proposes the following hypotheses:

H12: Athletes’ perceived behavioral control significantly influences their intention to receive the COVID-19 vaccine booster.

H13a: Athletes’ perceived behavioral control is a mediator between subjective norm and intention to receive the COVID-19 vaccine booster.

H13a: Athletes’ perceived behavioral control is a mediator between knowledge and intention to receive the COVID-19 vaccine booster.

#### 2.2.3 Response: Intention to Receive the COVID-19 Vaccine Booster

The intention to receive the COVID-19 vaccine booster has been a prevalent issue in recent years due to studies showing that antibodies would decline over time post-vaccination (Shrotri et al., 2021). The vaccine booster is one of the effective ways to maintain individual and public health. Thus, many studies focus on the factors influencing the intention to receive the COVID-19 vaccine booster. Such as pandemic fatigue [63], preventive practices [63], attitude [42, 64], subjective norms [42, 43], perceived behavioral control [42, 43], risk perception [42], vaccine hesitancy [42, 64], trust [64], social network [65], knowledge [65], and perceived vulnerability [43]. However, an integrating model is lacking to comprehend the intention to receive the COVID-19 vaccine booster, especially athletes. Due to athletes’ frequent interactions with diverse groups during training and competitions, they are at higher risk of contracting COVID-19. Thus, the primary aim of this study is to combine the TPB and SOR models to understand the factors affecting athletes’ intention to receive the COVID-19 vaccine booster.

#### 2.2.4 Moderator: Motivation

Motivation is crucial moderator in the integrating model, as it represents an individual’s internal drive or state that facilitates human behavior [66]. High motivation can trigger positively and actively emotional reflection. Motivation can be considered as a mediator to enhance the process of external stimuli to internal emotional responses, increase the likelihood of behavioral outcomes [67, 68]. Moderating effect of motivation is widely applied in management studies to examine employee behaviors. However, in the context of vaccination, no studies have used motivation as a moderator to explore its effects on psychological responses. Accordingly, the study employs motivation as a moderator to test the enhancing effect between athletes’ external stimuli to psychological reactions. The study proposes the following hypotheses:

H14a: Athletes’ motivation is a moderator between subjective norm and attitude.

H14b: Athletes’ motivation is a moderator between subjective norm and comment.

H14c: Athletes’ motivation is a moderator between knowledge and perceived behavioral control.

Based on an extensive literature review, the research framework is visually depicted in Figure 1.

**Figure 1.**
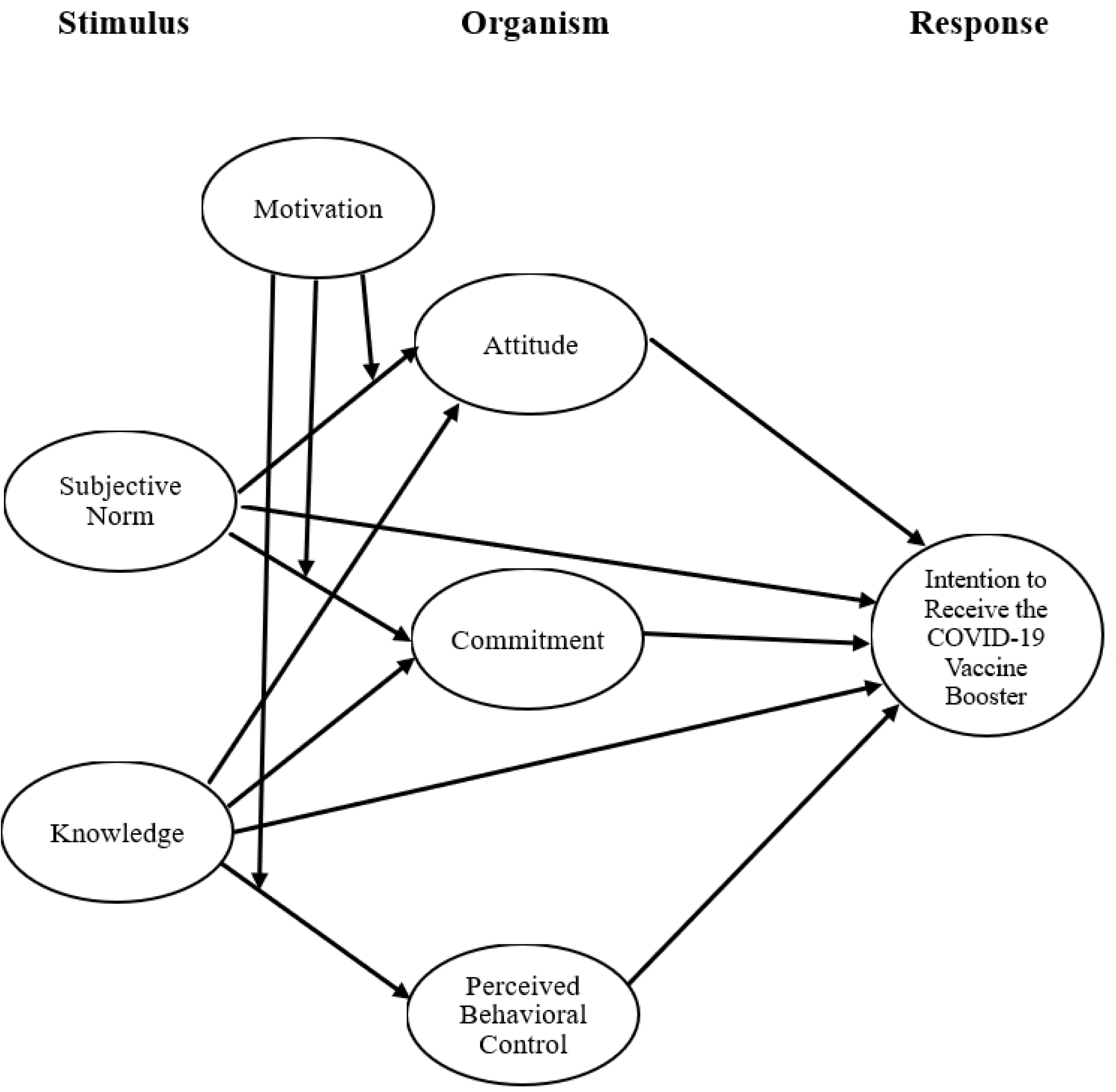
The fra1nework of integrating SOP and **TPB** 1nodel

## 3. Methods

### 3.1 Research Design

The primary purpose of this study was to use the TPB and SOR integrating model to identify the factors influencing athletes’ intention to receive the COVID-19 vaccine booster in Mainland China. The present study was conducted with a quantitative approach. A questionnaire was developed based on relevant previous studies. The study respondents were recruited from the 2021 National Games of the People’s Republic of China. Data collection was conducted through the administration of an online questionnaire. The collected data was analyzed by structural equation modeling (SEM).

### 3.2 Respondents

Purposive sampling was employed to select the athletes who are members of varsity sports teams and have experience in national games in Mainland China. The study excluded the athletes who have no experience in national games. Data collection was using a questionnaire survey, which took place during the 2021 National Games of the People’s Republic of China, held in Shaanxi from September 15 to September 27. To ensure adequate data collection, the study sought the cooperation with coaches who assisted in distributing the questionnaires to various sports groups of athletes, including soccer, bodybuilding, taekwondo, swimming, basketball, volleyball, table tennis, badminton, track and field, gymnastics, and traditional martial arts. A total of one thousand and fifty questionnaires were distributed. Consequently, sixty-nine questionnaires were identified as invalid and were excluded due to consistent or repetitive responses and incompleteness. Finally, nine hundred and eighty-one valid questionnaires were gathered, and the valid response rate was 93.4%.

### 3.3 Measurement

The questionnaire consisted of two sections. The first section is the scale, and the second section is respondents’ demographic information. In the first section, the items of the scale were developed following Ajzen (16) definitions of attitudes (5 items), subjective norm (4 items), perceived behavioral control (3 items), and intention to receive COVID-19 vaccine booster (3 items). Furthermore, the items of motivation (5 items) were developed based on Jang, Bai (66) definition of motivation. The commitment items (3 items) were built according to Lokhorst, Werner (58) opinions of commitment. Finally, The items of knowledge (4 items) were established by Jairoun, Al-Hemyari (49) definition (Table 1). This study used a Likert 7-point scale as the standard measurement.

**Table 1.**
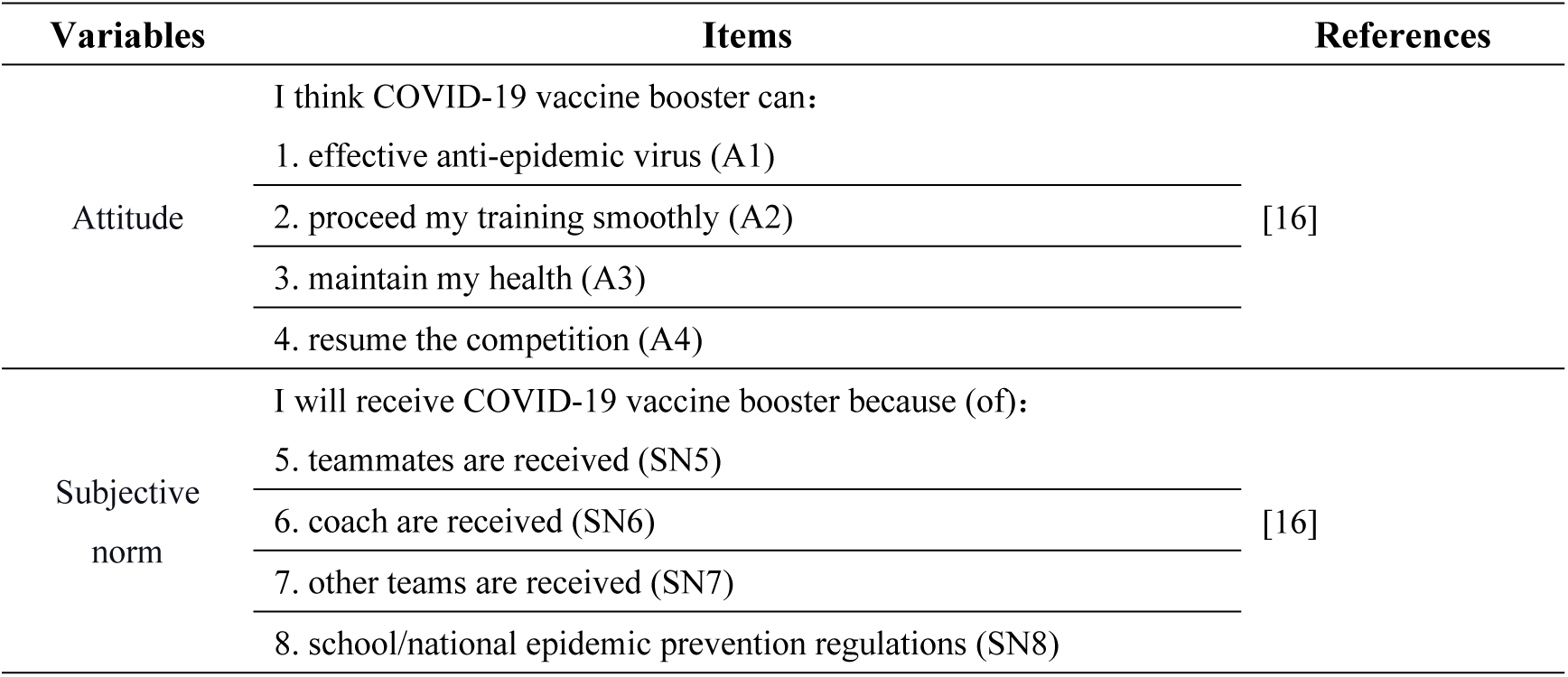
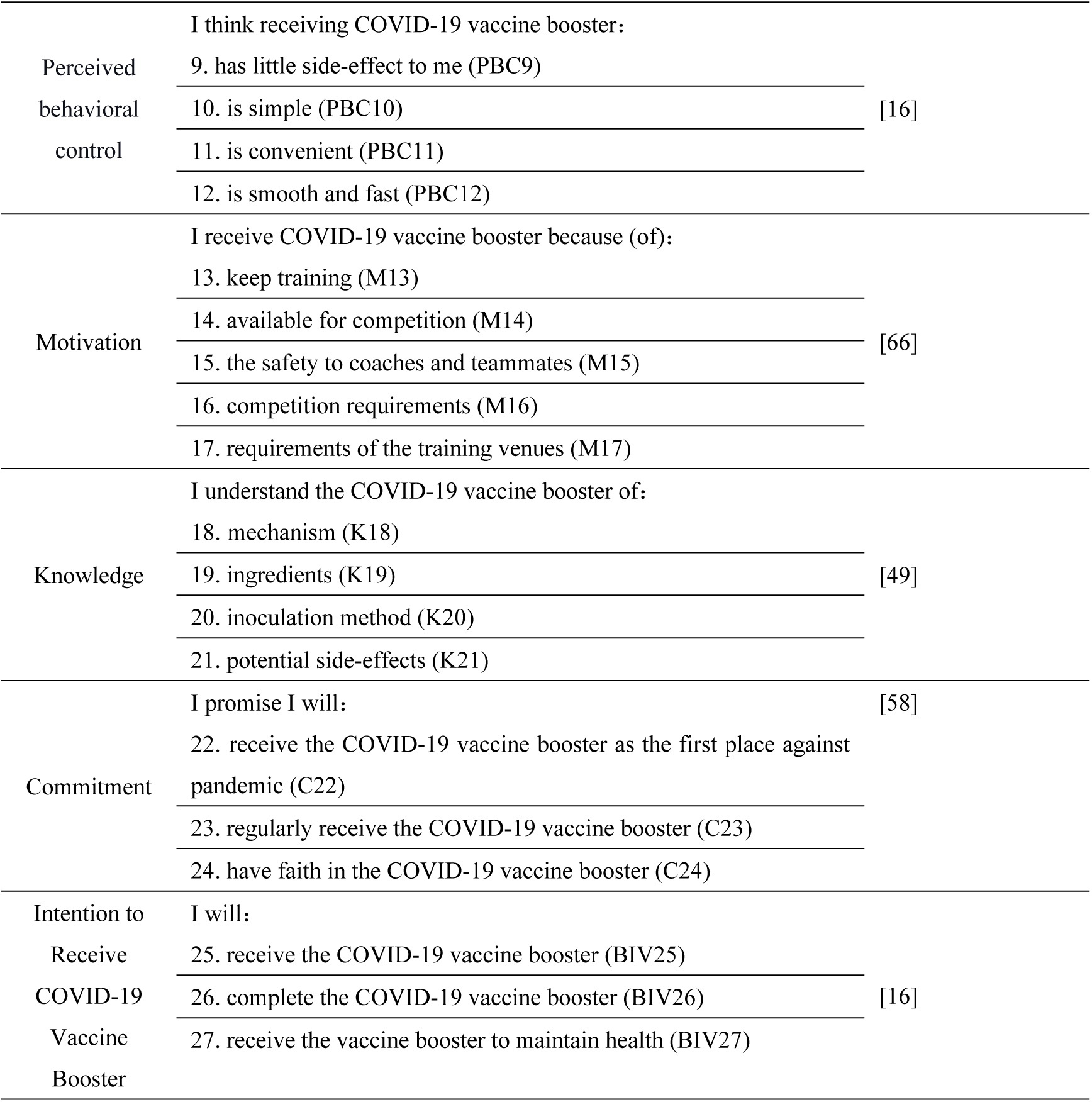
The items of the scale.

The second section of the questionnaire was used to collect respondents’ demographic information, including gender, grade level, major, sports expertise, place of residence, number of training days per week, training frequency per day, and duration of day training.

### 3.4 Ethical Statement

The present study was approved by the School Physical Science of Lingnan Normal University Review Board for the ethical approval (IRB# LNU2023112). Athletes participated in this study with guidance provided by researchers. Before distributing the questionnaires, the researchers explained the study’s objectives and the respondents’ rights, informed consent, to the coaches and athletes. The study employed an anonymous questionnaire survey to ensure that the respondents’ personal information was confidential. Respondents can refuse to answer questions or withdraw from the survey at any time. These key points were highlighted in the informed consent to achieve ethics standards.

### 3.5 Data Analysis

Statistical analyses for this study were conducted using SPSS 22.0 software and Smart-PLS 3.0 software. Descriptive analysis was employed to analyze respondents’ demographic information. Partial least squares structural equation modeling was used to analyze the reliability and validity, and verify the relationships in the integrating model.

## 4. Results and Discussion

### 4.1 Demographic

The respondents’ demographic information reveals that female athletes (54.3%) are slightly more than male athletes (45.7%). More than half of respondents’ majors are non-physical education (61.5%). Approximately 50.7% of the athletes specialize in taekwondo. 24.8% of the athletes train once a week, and 57.6% spend 61 to 120 minutes on daily training (Table 2).

**Table 2.** The respondents’ demographic (n=981)

| Variables | Frequency | Percentage (%) | Variables | Frequency | Percentage (%) |
| --- | --- | --- | --- | --- | --- |
| **Gender** |  |  | **Training days per week** |  |  |
| Male | 448 | 45.7 | One day | 243 | 24.8 |
| Female | 533 | 54.3 | Two days | 102 | 10.4 |
| **Major** |  |  | Three days | 137 | 14.0 |
| Physical education | 378 | 38.5 | Four days | 97 | 9.9 |
| Non-physical education | 603 | 61.5 | Five days | 173 | 17.6 |
| **Sports major** |  |  | More than six days | 229 | 23.3 |
| Soccer | 45 | 4.6 | **Duration of day training** |  |  |
| Aerobics | 41 | 4.2 | Under 60 minutes | 187 | 19.1 |
| Taekwondo | 496 | 50.7 | 61-120 minutes | 565 | 57.6 |
| Swim | 9 | 0.9 | 121-180 minutes | 174 | 17.7 |
| Basketball | 85 | 8.7 | 181 minutes or more | 55 | 5.6 |
| Volleyball | 99 | 10.1 |  |  |  |
| Table tennis | 20 | 2.0 |  |  |  |
| Badminton | 47 | 4.8 |  |  |  |
| Track and field | 26 | 2.7 |  |  |  |
| Gymnastics | 10 | 1.0 |  |  |  |
| Traditional martial arts | 23 | 2.3 |  |  |  |
| Others | 78 | 8.0 |  |  |  |

### 4.2 Reliability and Validity

The reliability and validity of the present study achieve the standard requirements. First, the observed variables’ factor loadings (FL) exceed 0.700 (range from 0.757 to 0.960), representing a high correlation between the observed and latent variables and great convergent validity. Second, the Cronbach’s alpha coefficient of the latent variables exceeds 0.900 (range from 0.901 to 0.949), which indicates strong internal consistency reliability. Third, the latent variables’ construction reliability (CR) coefficients exceed 0.800 (range from 0.925 to 0.961), showing substantial correlations between the observed variables. Fourth, the average extracted variance (AVE) of the variables surpasses the standard values of 0.500 (range from 0.755 to 0.884), which indicates that the observed variables can explain each latent variable over 50% of the variance (Table 3). Finally, the square root of the AVE of each latent variable is higher than the correlation coefficients between the variables, demonstrating that the potential variables possess great discriminant validity. The integrating model achieves strong reliability and validity (Table 4).

**Table 3.** Reliability and validity.

| Variables | Items | FL | M | Cronbach's $\alpha$ | CR | AVE |
| --- | --- | --- | --- | --- | --- | --- |
| Attitude | A1 | 0.942 | 6.606 | 0.946 | 0.961 | 0.862 |
|  | A2 | 0.952 |  |  |  |  |
|  | A3 | 0.920 |  |  |  |  |
|  | A4 | 0.898 |  |  |  |  |
| Subject norm | SN5 | 0.898 | 6.074 | 0.901 | 0.925 | 0.755 |
|  | SN6 | 0.908 |  |  |  |  |
|  | SN7 | 0.904 |  |  |  |  |
|  | SN8 | 0.757 |  |  |  |  |
| Perceived behavioral control | PCB9 | 0.943 | 6.229 | 0.918 | 0.948 | 0.860 |
|  | PCB10 | 0.908 |  |  |  |  |
|  | PCB11 | 0.929 |  |  |  |  |
|  | PCB12 | 0.943 |  |  |  |  |
| Motivation | M13 | 0.896 | 6.480 | 0.949 | 0.961 | 0.831 |
|  | M14 | 0.918 |  |  |  |  |
|  | M15 | 0.884 |  |  |  |  |
|  | M16 | 0.936 |  |  |  |  |
|  | M17 | 0.923 |  |  |  |  |
| Knowledge | K18 | 0.910 | 6.149 | 0.901 | 0.930 | 0.770 |
|  | K19 | 0.880 |  |  |  |  |
|  | K20 | 0.887 |  |  |  |  |
|  | K21 | 0.830 |  |  |  |  |
| Commitment | C24 | 0.929 | 6.552 | 0.913 | 0.945 | 0.852 |
|  | C25 | 0.946 |  |  |  |  |
|  | C26 | 0.894 |  |  |  |  |
| Intention to Receive COVID-19 Vaccine Booster | IB22 | 0.960 | 6.628 | 0.934 | 0.958 | 0.884 |
|  | IB23 | 0.905 |  |  |  |  |
|  | IB27 | 0.954 |  |  |  |  |

Note: FL=Factor loading; M=Mean; CR= Construction reliability; AVE= Average extraction variance

**Table 4.** Discriminant analysis.

|  | A | C | BI | K | M | PBC | SN |
| --- | --- | --- | --- | --- | --- | --- | --- |
| A | **0.928** |  |  |  |  |  |  |
| C | 0.857 | **0.923** |  |  |  |  |  |
| BI | 0.824 | 0.902 | **0.940** |  |  |  |  |
| K | 0.684 | 0.698 | 0.638 | **0.877** |  |  |  |
| M | 0.780 | 0.739 | 0.733 | 0.612 | **0.912** |  |  |
| PBC | 0.800 | 0.782 | 0.774 | 0.690 | 0.737 | **0.927** |  |
| SN | 0.649 | 0.632 | 0.603 | 0.625 | 0.631 | 0.614 | **0.869** |

Note: A= Attitude; C= Commitment; BI= Behavioral control; K= Knowledge; M=Motivation; PBC= Perceived behavioral control; SN= Subject norm

### 4.3 Model Fit

Goodness-of-fit (GoF) is the essential model evaluation index for PLS-SEM. Akter, D’ambra (69) suggest that if the GoF is higher than 0.360, it means that the model has a high level of model fit; between 0.250 to 0.350 is a medium level of model fit; between 0.10 to 0.240 is acceptable; below 0.100 means the model fit is unacceptable. The formula of GoF is as follows:

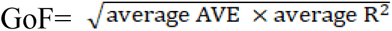

The GoF of this study is calculated to be 0.791, which means that the integrating model has a high level of model fit.

### 4.4 Structural Model Analysis

The results of the structural model analysis (Figure 2) indicate that stimulus to organism and response, the athletes’ subjective norm significantly influences attitude (β=0.342*, p<0.05) and commitment (β=0.304*, p<0.05) but does not significantly influence the intention to receive COVID-19 vaccine booster (β=0.008, p>0.05). On the other side, the athletes’ knowledge of the COVID-19 vaccine booster has a significant effect on attitude (β=0.472*, p<0.05), commitment (β=0.510*, p<0.05), and perceived behavioral control (β=0.690*, p<0.05). However, knowledge does not significantly influence the athletes’ intention to receive the COVID-19 vaccine booster (β=0.049, p>0.05). Knowledge more substantially affects athletes’ attitudes and commitment to receiving the COVID-19 vaccine booster compared to subjective norm. Moreover, knowledge can effectively enhance athletes’ perceived behavioral control of getting the COVID-19 vaccine booster.

**Figure 2.**
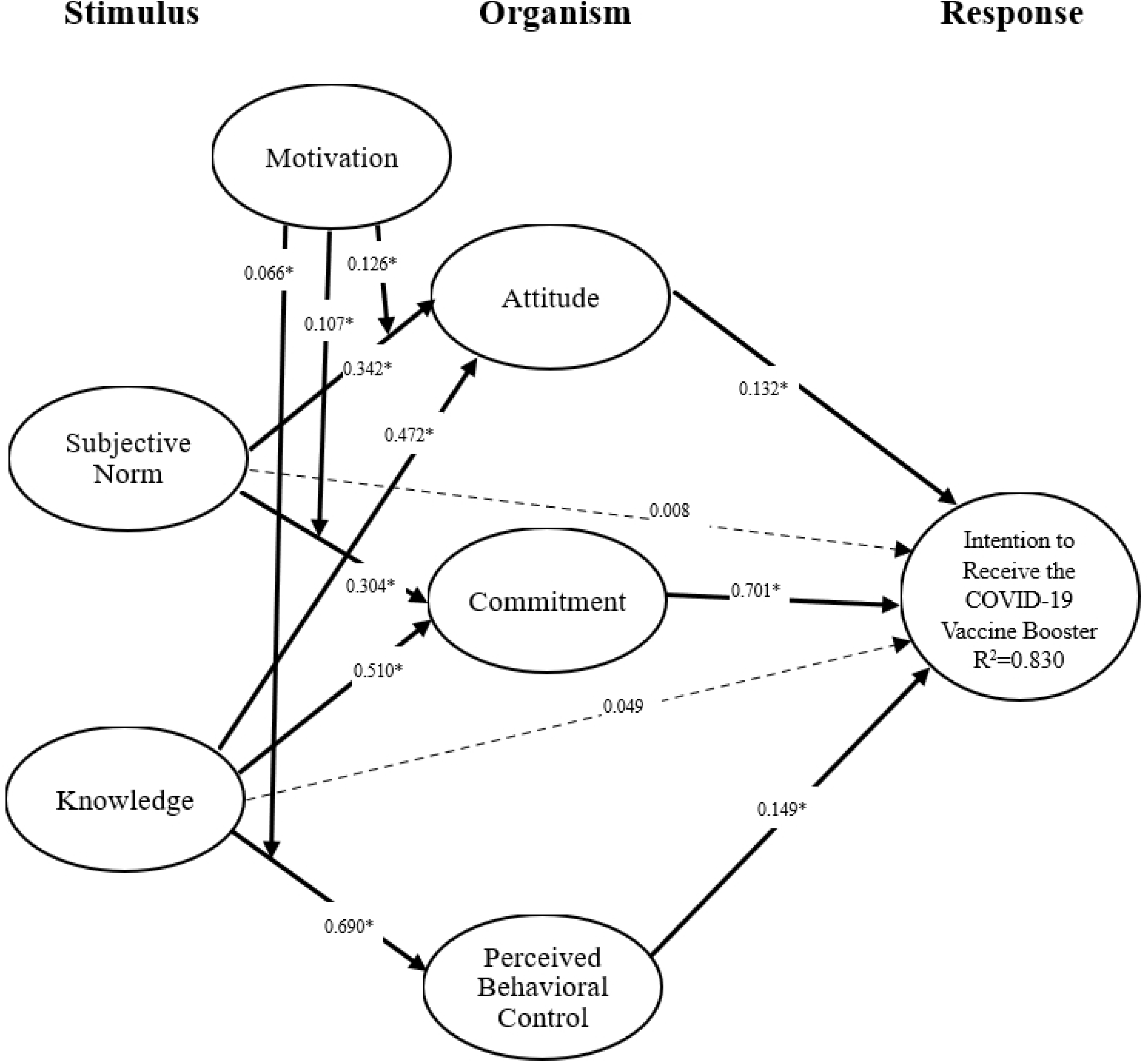
The results of integrating SOP and **TPB** model

Of the organism to response, athletes’ attitude (β=0.132*, p<0.05), commitment (β=0.701*, p<0.05), and perceived behavioral control (β=0.149*, p<0.05) significantly influence the intention to receive COVID-19 vaccine booster. Commitment is the most vital organism factor in promoting athletes’ intention to receive the COVID-19 vaccine booster. Furthermore, in the receiving COVID-19 vaccine booster context, attitude, commitment, and perceived behavioral control fully mediate the integrating model. Attitude has mediating effects between subjective norm (β=0.045*, p<0.05), knowledge (β=0.062*, p<0.05), and the intention to receive COVID-19 vaccine booster. Subjective norm (β=0.213*, p<0.05) and knowledge (β=0.358*, p<0.05) can indirectly influence the intention to receive the COVID-19 vaccine booster via commitment. Perceived behavioral control mediates knowledge (β=0.103*, p<0.05) and the intention to receive the COVID-19 vaccine booster. Accordingly, knowledge to commitment is the most effective path to increase the athletes’ intention to receive COVID-19 vaccine booster.

Furthermore, the athletes’ motivation can moderate the relationships between subjective norm and attitude (β=0.126*, p<0.05), subjective norm and commitment (β=0.107*, p<0.05), as well as knowledge and perceived behavioral control (β=0.066*, p<0.05). Subjective norm interaction with motivation can enhance athletes’ attitude and commitment. Athletes’ perceived behavioral control can be improved by knowledge-motivation interaction. The integrating model can explain 83% of athletes’ intention to receive the COVID-19 vaccine booster.

## 5. Discussion

The present study combines the TPB and SOR to identify the factors influencing athletes’ intention to receive the COVID-19 vaccine booster, as well as the moderating effects of motivation in the model. Overall, the integrating model provides insight into why athletes receive COVID-19 vaccine booster. Subjective norm and knowledge are the vital external stimuli that positively trigger athletes’ attitude, commitment, and perceived behavioral control of getting the COVID-19 vaccine booster. Subjective norm can significantly influence individual emotional perceptions [45]. Such as Kim, Ham (46) and Shin and Hancer (47) they were pointed out that the crucial social group can positively or negatively shape individuals’ attitude towards a particular behavior. Commitment is produced because of individuals’ innate desire for social acceptance and cohesion to seek external validation and fear social repercussions [48]. Thus, to correspond to expectations within social circles, subjective norms from coaches and teammates are crucial to prompting athletes to receive the COVID-19 vaccine booster.

However, the study found that subjective norm and knowledge do not directly affect athletes’ intention to receive the COVID-19 vaccine booster. This finding differs from most related studies [42–44, 50]. Athletes often prioritize personal experiences and beliefs over societal and peer pressure because of their high self-awareness [70]. Thus, significant others only influence athletes’ internal emotions but do not directly affect their decision-making about receiving the COVID-19 vaccine booster. In addition, the concept of the SOR model refers to the response as the result of internal reflection triggered by external factors [38]. Thus, knowledge does not directly influence athletes’ intention to receive the COVID-19 vaccine booster. Instead, it must be internalized into positive attitudes, commitment, and perceived behavioral control, leading to behavioral intention.

Athletes’ attitude, commitment, and perceived behavioral control can increase their intention to receive the COVID-19 vaccine booster. The results echo related studies with similar findings [42, 53, 54, 59, 61]. Undoubtedly, positive attitude, strong commitment, and high perceived behavioral control are helpful to encourage athletes to overcome the barriers and engage in the intention to get the COVID-19 vaccine booster [16, 58, 59]. Commitment is the strongest predictor in the integrating model because its emotional and psychological attachment solidly connects with the specific behavioral intention [58, 59]. When athletes believe the COVID-19 vaccine booster is an effective way to maintain their health and continue their training, the belief will transfer into the commitment that improves their intention to receive the COVID-19 vaccine booster.

Furthermore, attitude, commitment, and perceived behavioral control can indirectly increase athletes’ intention to receive the COVID-19 vaccine booster. These organism factors play the full mediators in the model. These findings are similar to the previous studies [55–57, 71, 72]. In the present study, subjective norms and knowledge are treated as stimuli that arouse athletes’ internal reflections, including attitude, commitment, and perceived behavioral control, and further produce their behavioral intention. The SOR model argues that the organism is a crucial process in triggering the response because it is an inevitable mechanism [39]. Particularly, athletes’ commitment is the most significant mediator influencing the intention to receive the COVID-19 vaccine booster in the model. Commitment reflects individuals’ strong internal conviction and dedication that forces to act subsequent behaviors [73]. When athletes believe in the benefits of the COVID-19 vaccine booster, they are genuinely committed to receiving the vaccine. Knowledge is regarded a catalyst to foster athletes’ commitment. Li et al. (2021) argued that a lack of vaccine knowledge is the primary reason for vaccine hesitancy, and increasing this knowledge can bolster vaccine trust and commitment. Accordingly, enhancing athletes’ vaccine knowledge is the priority strategy to improve their intention.

Motivation can boost the relationship between stimulus and organism, including subjective norm and attitude, subjective norm and commitment, as well as knowledge and perceived behavioral control. Individuals with strong intrinsic motivation are more likely to interact with external stimuli, thereby increasing their willingness to perform specific behaviors [67, 68]. Thus, motivation amplifies the effect of athletes’ stimulus (subjective norm and knowledge) to the organism (attitude, commitment, and perceived behavioral control) in the model. Therefore, boosting athletes’ motivation for the COVID-19 vaccine booster is key to enhancing their attitude, commitment, and perceived behavioral control towards vaccination. Getting the COVID-19 vaccine booster should not just be mandatory; appropriate incentives are also essential.

## 6. Contributions and Implications

The present study provides several theoretical contributions and practical implications for improving athletes’ intention to receive the COVID-19 vaccine booster. The contributions and implications are interpreted as follows:

### 6.1 Theoretical Contributions

There are two significant theoretical contributions in the study. First, the study combines TPB and SOR model to understand athletes’ pattern of receiving the COVID-19 vaccine booster. The integrating model provides a comprehensive picture of the factors and paths influencing athletes’ intentions. The findings address the lack of an integrated model examining athletes’ intention to receive the COVID-19 vaccine booster. The study proves that the relationships between variables align with the SOR model. Also, the subjective norm is confirmed as an external factor, and attitude and perceived behavioral control are verified as internal reflections. The study suggests that the model can be used to explore the vaccinated behavior in different issues and populations in future study.

In addition, this study reveals that motivation is a moderating variable prompting athletes towards a positive psychological reflection toward the COVID-19 vaccine booster. Motivation is treated as an independent variable in public health issues to explore its effect on behavioral intention or psychological status. Usually, motivation is considered a moderator to understand its reinforcement on individual psychological responses in human resource management and business administration fields. However, this study confirms that motivation is an essential moderator in public health issues, which can positively influence attitudes, commitment, and subjective norms toward vaccines. Future studies on public health can consider motivation as a moderator to explore relevant issues.

### 6.2 Practical Implications

Here are two practical implications for promoting vaccine acceptance. First, eliminating and preventing the spread of inaccurate vaccine information is paramount. The correct knowledge is the first and most crucial step to increase athletes’ intention to receive COVID-19 vaccine booster. Salzburg Statement on Vaccination Acceptance suggests that (1) distinguishing levels of evidence in social media organizations and major search engines, (2) providing support from governments, policymakers, advocacy groups, educators, and philanthropists, as well as (3) educating by healthcare professionals and educators can improve individuals’ correct knowledge of the vaccine [74]. Therefore, Internet information and health education, and support from relevant stakeholders for athletes are crucial. Athletes should learn knowledge about public health from professionals during their off-training periods.

Second, incentives are necessary to increase the athletes’ motivation to get the COVID-19 vaccine booster, which can effectively enhance their positive attitude and trust towards the vaccine booster. For example, in the US, vaccination incentives include free beers, $100 savings bonds, scholarships, etc. Concurrently, delivering accurate vaccine information to the people establishes their long-term commitment and improves their received intention. Hence, the specific incentives combined with knowledge should be designed for athletes, such as offering free training equipment to those who receive the vaccine education and the COVID-19 vaccine booster. It can boost athletes’ vaccination intentions and substantially enhance their attitudes and commitment.

## 7. Limitations of the Study

This study was conducted rigorously to comprehend athletes’ intention to receive the COVID-19 vaccine booster. Nevertheless, there are several study limitations. Firstly, the respondents in the study were amateur athletes in Mainland China. The study results cannot be generalized to other populations. The future study could consider other populations, i.e., professional athletes, to compare the differences in the intention to get the COVID-19 vaccine booster. Additionally, this study was conducted in Mainland China. Thus, the findings are only applicable to Chinese athletes. Due to cultural differences, the results cannot be generalized to athletes from other countries. Future research can include athletes from other countries to test if similar results are observed. Third, this study utilized a quantitative approach to verify the relationships in the model. However, the reasons behind forming relationships are ambiguous. Therefore, future research can consider incorporating qualitative methods to provide detailed information explaining the athletes’ intention to receive the COVID-19 vaccine booster.

## 8. Conclusions

This study proved that integrating TPB and SOR model can effectively explain athletes’ intention to receive the COVID-19 vaccine booster. Subjective norm and knowledge significantly affect attitude, commitment, and perceived behavioral control. Attitude, commitment, and perceived behavioral control influence athletes’ intention to receive the COVID-19 vaccine booster and serve as full mediators in the model. The path from knowledge to commitment greatly affects athletes’ intention. Moreover, motivation significantly moderates the model’s relationship between stimulus and organism. High motivation can amplify athletes’ internal responses when interacting with external factors.

## Data Availability

All relevant data are within the manuscript and its Supporting Information files.

